# Understanding the Impact of Comorbidity-Interaction in Patients Undergoing Transcatheter Edge-to-Edge Mitral Valve Repair on Outcomes

**DOI:** 10.1101/2023.04.14.23288606

**Authors:** Ankit Agrawal, Shivabalan Kathavarayan Ramu, Shashank Shekhar, Toshiaki Isogai, Agam Bansal, James Yun, Grant W. Reed, Rishi Puri, Amar Krishnaswamy, Samir R. Kapadia

**Affiliations:** Department of Cardiovascular Medicine, Heart, Vascular and Thoracic Institute, Cleveland Clinic, Cleveland, OH 44195, USA; Department of Thoracic and Cardiovascular Surgery, Heart, Vascular and Thoracic Institute, Cleveland Clinic, Cleveland, Ohio, USA

**Keywords:** M-TEER, Non-cardiac comorbidities, MACE, MitraClip

## Abstract

**BACKGROUND:** Transcatheter Edge-to-Edge Mitral Valve Repair (M-TEER) is an accepted procedure for high-risk surgical patients with degenerative and functional mitral regurgitation. Non-cardiovascular comorbidities (NCCs) are highly prevalent in patients undergoing M-TEER. Although the impact of mitral valve anatomy and cardiac comorbidities in determination of M-TEER outcomes has been studied, precise understanding of the effect of the burden of NCCs on patients undergoing M-TEER remains unclear for acute outcomes. Our objective was to identify the association of NCC comorbidity-interaction patterns in patients undergoing M-TEER on length of stay (LOS), cost of care, and in-hospital major adverse cardiovascular events (MACE).

**METHODS:** 9 245 admissions from the Nationwide Readmission Database that underwent M-TEER between 2015 and 2018 were included in the study. Patients were categorized by the overall burden of non-cardiovascular comorbidities (0, 1, 2, and ≥ 3). NCC included chronic liver disease, chronic lung disease, obesity, diabetes mellitus, dementia, major depressive disorder, chronic anemia, chronic kidney disease including end-stage renal disease (ESRD) on dialysis, and malignancy. Logistic Regression and Machine Learning (ML) algorithms were used to assess associations between comorbidity burden and in-hospital MACE.

**RESULTS:** Out of 9 245 index admissions, in-hospital MACE was recorded in a total of 504 (5.3 %). Of these, the majority (30.4%) had one NCC (n = 2 861). Patients with at least three NCCs had the longest median LOS [3.0, IQR (1.0 – 11.0)] and highest median cost of hospital care [$47 275, IQR (34 175.8 – 71 149.4)]. The Gradient Boosting (GB) classifier performed the best in predicting MACE with an AUROC of 96 % (95% CI: 0.95 – 0.97). The top features of importance that predicted in-hospital MACE were admission type, number of NCCs, and age in descending order.

**CONCLUSIONS:** Calibrated GB classifier identified patients with three NCCs as the subset of admission having the highest probability of a positive MACE outcome.

## INTRODUCTION

The incidence of valvular HD is 64 per 100 000 person-years in the United States (U.S.), with almost 24.2% of all valve diagnoses being mitral regurgitation (MR). Around 16 250 people in the U.S. have MR secondary to ischemia or left ventricular failure (type IIIb MR).^1^ The Endovascular Valve Edge-to-Edge Repair Study (EVEREST II) compared treatment with the percutaneous MitraClip (MC) (Abbott Vascular-Structural Heart, Menlo Park, CA) device with surgical mitral valve repair. It showed a superior safety of M-TEER compared to the surgical mitral valve group, even though the M-TEER group had a higher comorbidity burden.^2^ But even with both the groups combined, many patients did not meet the threshold of a Society of Thoracic Surgeons score of ≥ 12. However, in real-world settings, patients who were chosen for the M-TEER procedure have a different comorbidity phenotype. It has been shown that, in patients with a Charlson comorbidity index of ≥ 2, M-TEER has a lesser incidence of inpatient complications compared to surgical repair.^3^ Still, though the impact of diabetes mellitus,^4^ chronic obstructive lung disease,^5^ liver cirrhosis,^6^ and chronic kidney disease^7^ are known, enough data on the cumulative burden of non-cardiac comorbidities (NCCs) on the M-TEER outcomes and hospital cost, is lacking. An intuitive knowledge of the impact of noncardiovascular comorbidities on M-TEER outcomes will further improve patient outcomes by assisting in patient care and direct hospital resource management.

In an observational study done on two hundred and sixty-four patients from the EVEREST II study, patients with atrial fibrillation have been shown to have more medical non-cardiovascular comorbidities and need more after-load-reducing medications.^8^ In an analysis that investigated the influence of NCC on outcomes of patients enrolled in the German transcatheter mitral valve interventions (TRAMI) registry, it was shown that patients with multiple NCCs more often needed intensive care, had a longer hospital stay, and had higher rates of rehospitalizations.^9^ Little is known about the impact of the cumulative burden of NCCs on outcomes following M-TEER.

Given the current picture, studying the burden of NCCs in M-TEER patients is imperative. Additionally, though a linear analysis might help understand the burden of NCC on M-TEER outcomes, it will fail to reveal non-linear relationships between clinical features and outcomes that help to recognize patient microclusters with different probabilities of a positive MACE outcome. Data which gives a nationwide perspective of the same is limited. Therefore, we aim to evaluate the influence of select NCCs and their interaction effects on major adverse cardiovascular events (MACE) outcomes following M-TEER from a nationwide population-based registry. We have also compared the linear analysis methods with machine learning methods to elucidate the non-linear relationships between NCC and other features that govern a positive MACE outcome.

## METHODS

### DATA SOURCE

We used the 2015-2018 Nationwide Readmissions Database (NRD). NRD is a national publicly available database of all-payer hospital inpatient stays in the U.S. sponsored by the Agency for Healthcare Research and Quality as part of the Healthcare Cost and Utilization Project (HCUP). It is collected from twenty eight states across the U.S. that are geographically dispersed. The NRD accounts for 59.7% of the total U.S. resident population and 58.7% of all hospitalizations across the U.S. Demographics, comorbidities, expected payment source, total charges and hospital cost, length of stay, inpatient procedures, in-hospital mortality, and post-discharge readmissions.^10^ As the NRD contains deidentified patient information, this study was exempt from review by the Cleveland Clinic Institutional Review Board.

### STUDY POPULATION

We used International Classification of Diseases (ICD)-09-CM codes and ICD-10-CM codes to identify variables from the NRD. Hospital stays for patients aged ≥ 18 years who underwent M-TEER were only included. NCCs evaluated were chronic liver disease, chronic lung disease, obesity, diabetes mellitus, dementia, major depressive disorder, chronic anemia, end-stage renal disease (ESRD) on dialysis, chronic kidney disease, and malignancy.

### PRIMARY OUTCOMES

The primary outcome of interest included identifying variables that predict MACE in M-TEER patients and constructing explainable Machine Learning (ML) models to measure the impact of the burden of NCC on MACE. The study flow is summarized in Figure 1. Details of the data processing, model selection, training and validation are available in the Supplemental Methods.

**Figure 1.**
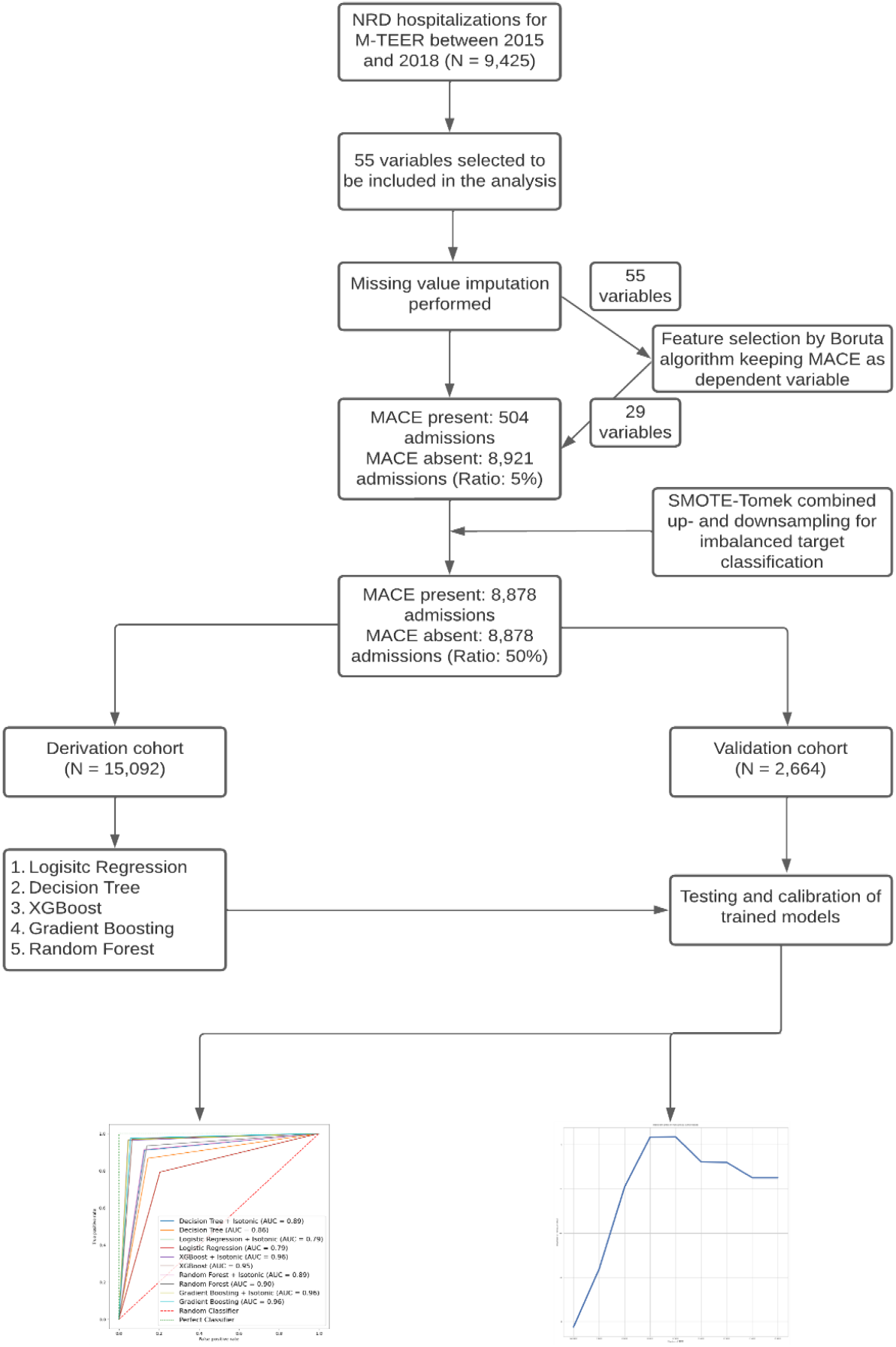
Study-flow showing the sequence of steps involved.

### STATISTICAL ANALYSIS

Kolmogorov-Smirnov Z test was performed to test whether the variables were normally distributed according to various grouping factors. Kruskal-Wallis test was conducted for each variable to determine if differences in the admission characteristics between risk groups were significant. Descriptive statistics were used to summarize the percentages (for categorical variables) and means (± standard error (SE)) (for continuous data). Post hoc analyses for between–category comparisons in each NCC group were also performed and adjusted for multiple comparisons using the Bonferroni method. If Kruskal-Wallis test has been performed to test a variable, post hoc Tukey-Kramer test was conducted. Statistical analyses were performed using SPSS ver. 23.0 (IBM Corp., USA).^20^ Feature selection was performed in R (version 4.2.2; R Project for Statistical Computing) using Boruta package.^21^ Strength of association between features selected by Boruta is calculated for nominal vs. nominal with a bias-corrected Cramer’s V, numeric vs. numeric with Spearman correlation, and nominal vs. numeric with one-way analysis of variance (ANOVA). Machine learning classifiers were developed in Python 3.8.9 programming language using scikit-learn library.^22^ Statistical significance was defined as two-tailed p-value less than 0.05. We adhered to the recommendations for reporting machine learning analyses in clinical research.^23^

## RESULTS

### COHORT ANALYSIS BY BURDEN OF NCCs

Out of the total 9 425 admissions with history of M-TEER between 2015-18 identified in the NRD, 504 (5.3%) had encountered MACE in-hospital. Table 1 provides the characteristics of admissions included in the study. Of these, the majority (30.4%) had one NCC (2 861), 2 405 (25.5%) had no NCC, 2 189 (23.2%) had two NCC, and 1 970 (21%) had at least three NCCs. The median age of patients was highest in admissions with no NCC [83.0, IQR (76 – 87)] and lowest in admissions with atleast three NCCs [77.0, IQR (69 – 83)] (*P* < 0.001). Among M-TEER admissions, number of NCCs (collapsed into 4 ordered categories) and sex-based difference was significantly associated, *P* < 0.001. Post hoc comparisons of sex categories revealed that a higher proportion of females had no NCCs. In comparison, sex-based difference was statistically similar among those with atleast one NCCs with more proportion of males. Mean Elixhauser score was highest in admissions with atleast three NCCs (7.5 ± 6.69) and lowest in admissions with no NCC (1.4 ± 2.98), as expected. A post hoc Tukey test showed that the mean Elixhauser Score differed significantly between NCC categories at *P* < 0.05. Medicare was the largest payer in all the categories (*P* < 0.001). The majority of admissions happened in rural hospitals (*P* < 0.001). Most of the admissions with no or one NCC happened in high-volume centers (*P* < 0.05) whereas, majority of patients with two NCCs got admitted in low-volume centers (*P* < 0.05). The majority of discharges to skilled nursing facility (SNF) happened in admissions with atleast three NCCs, and the least number of patients with no NCCs got discharged to SNF (*P* < 0.001). The median cost was the highest in admissions with atleast three NCCs amounting to $47 275.0 [IQR (34 175.8 – 71 149.4)] and lowest in admissions with no NCCs amounting to $39 374.3 [IQR (30 200.53 – 52 443.03)]. A post hoc Tukey after ANOVA revealed that the differences between the categories of NCCs are significant (*P* < 0.05) except between admissions with no NCC and admission with one NCC (*P* = 0.08). With every addition of one comorbidity in admission with one NCC, the mean hospital cost increases by atleast $6 400 (Table 1).

**Table 1.**
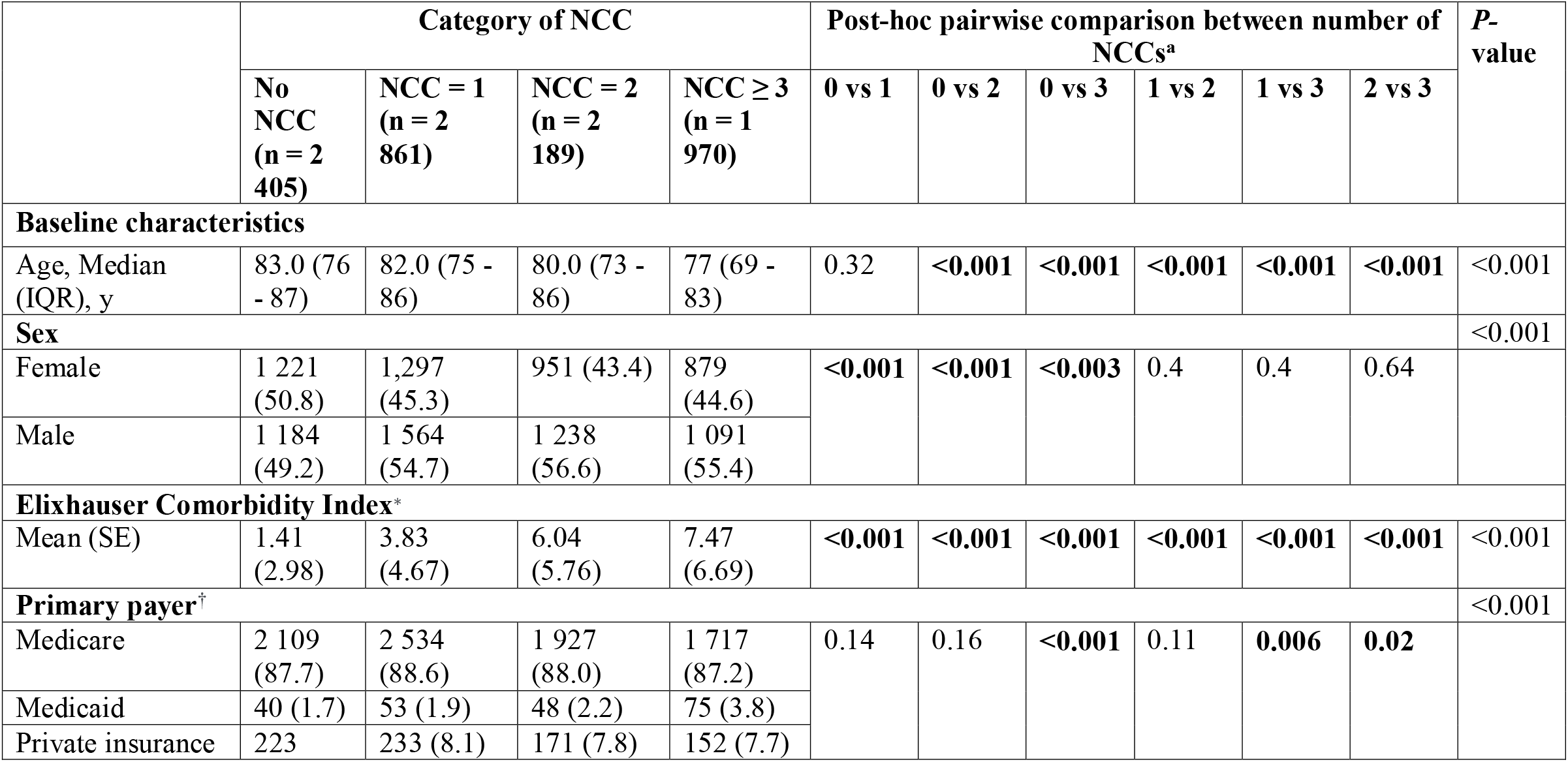
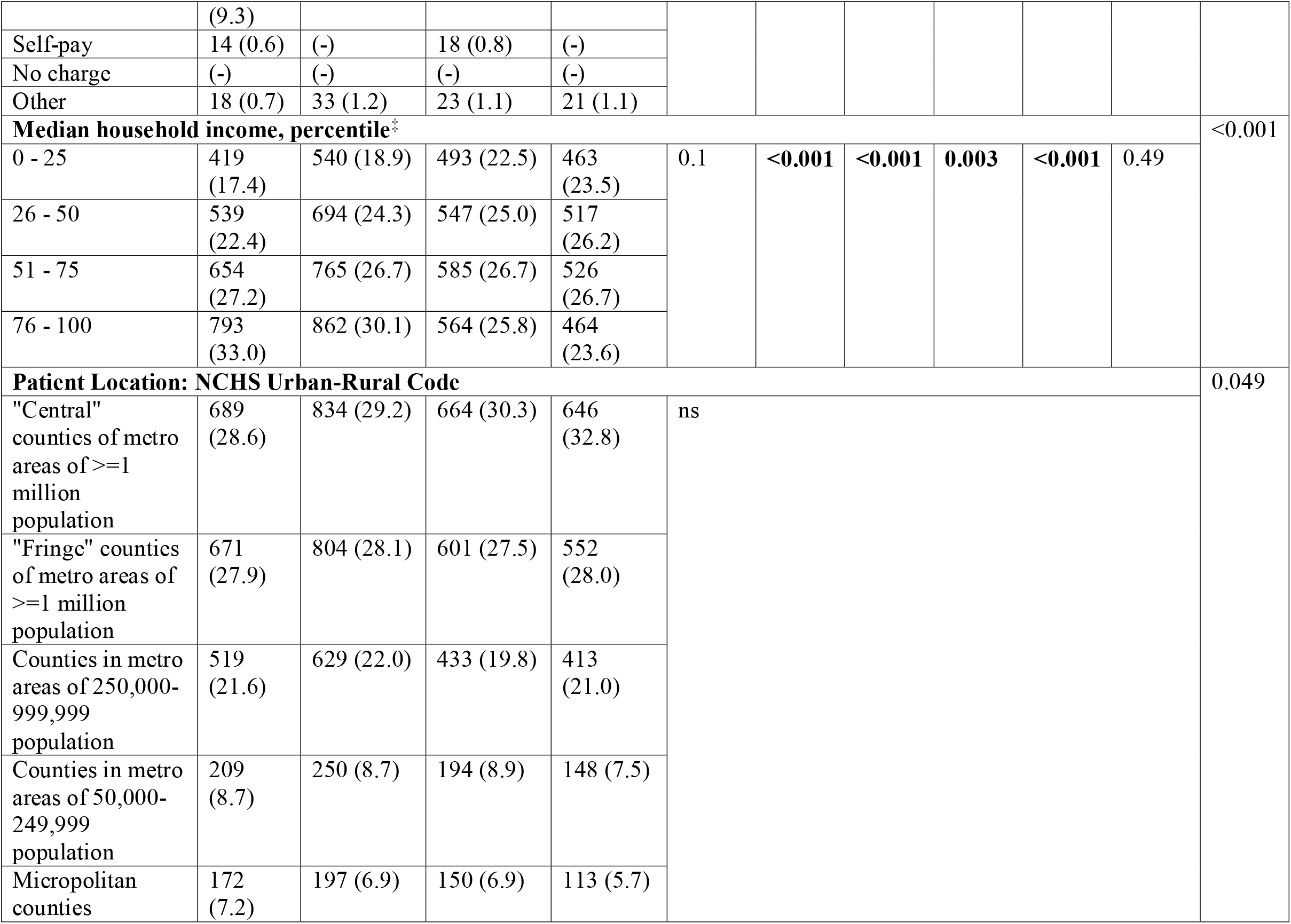
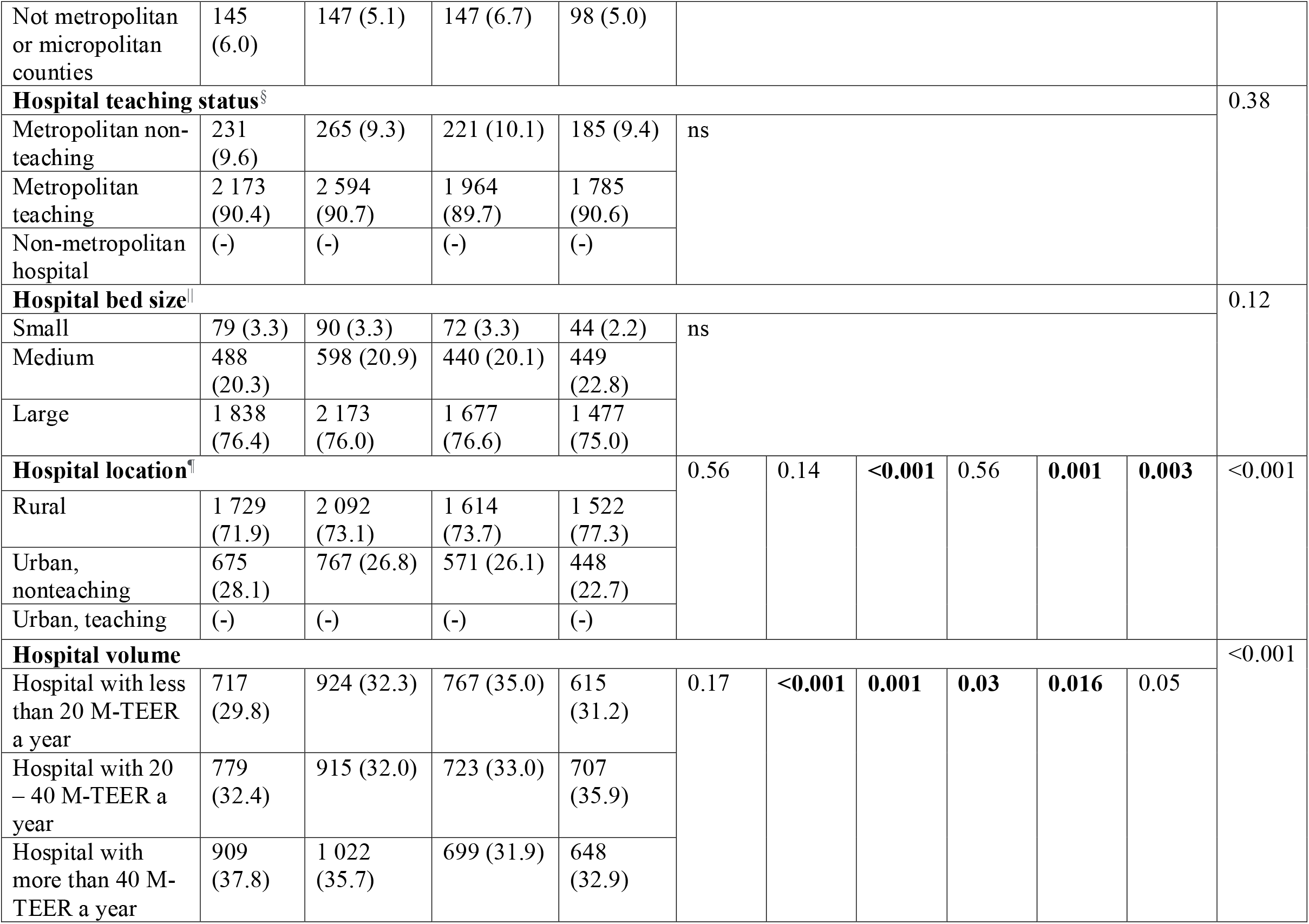
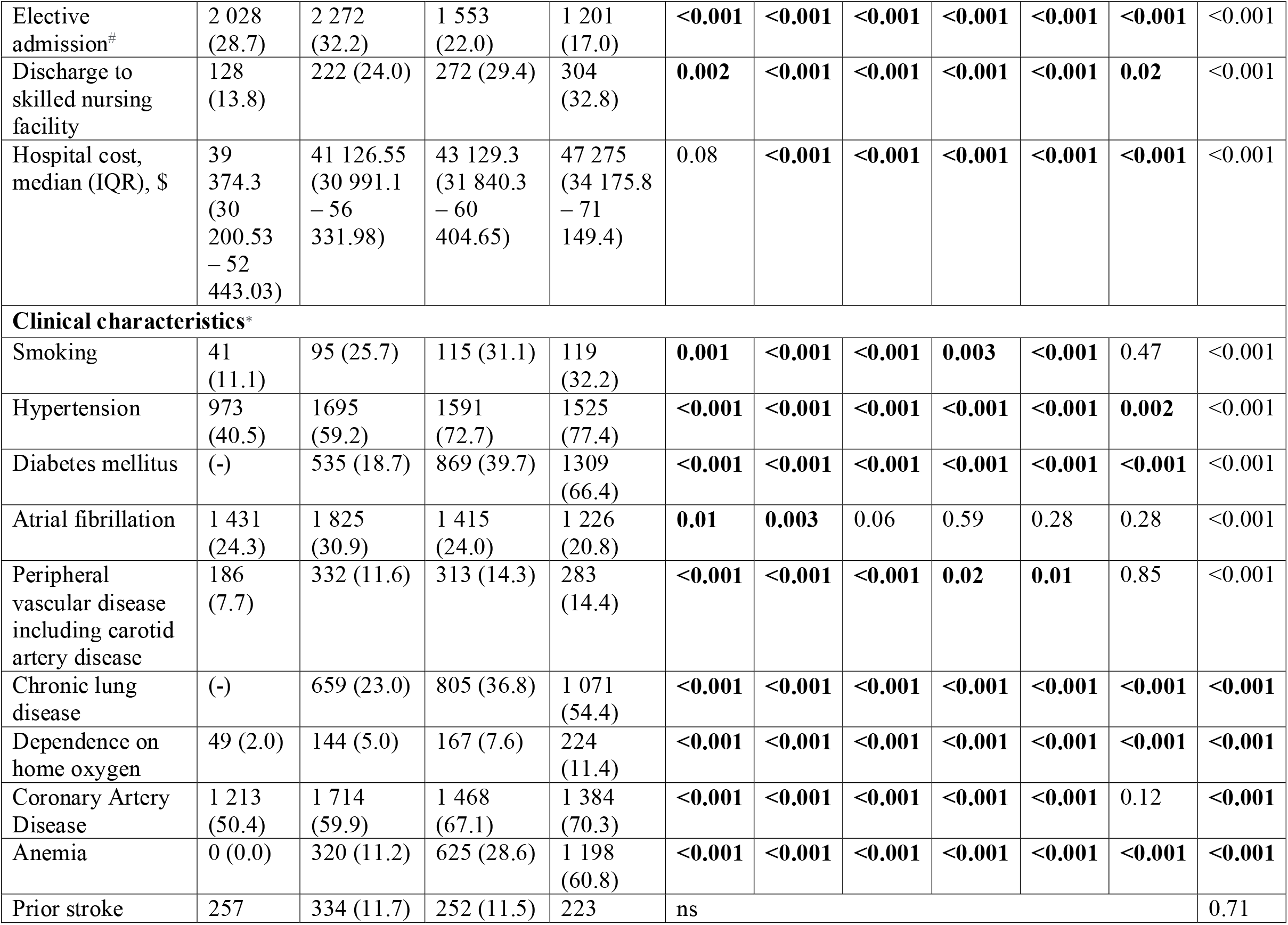
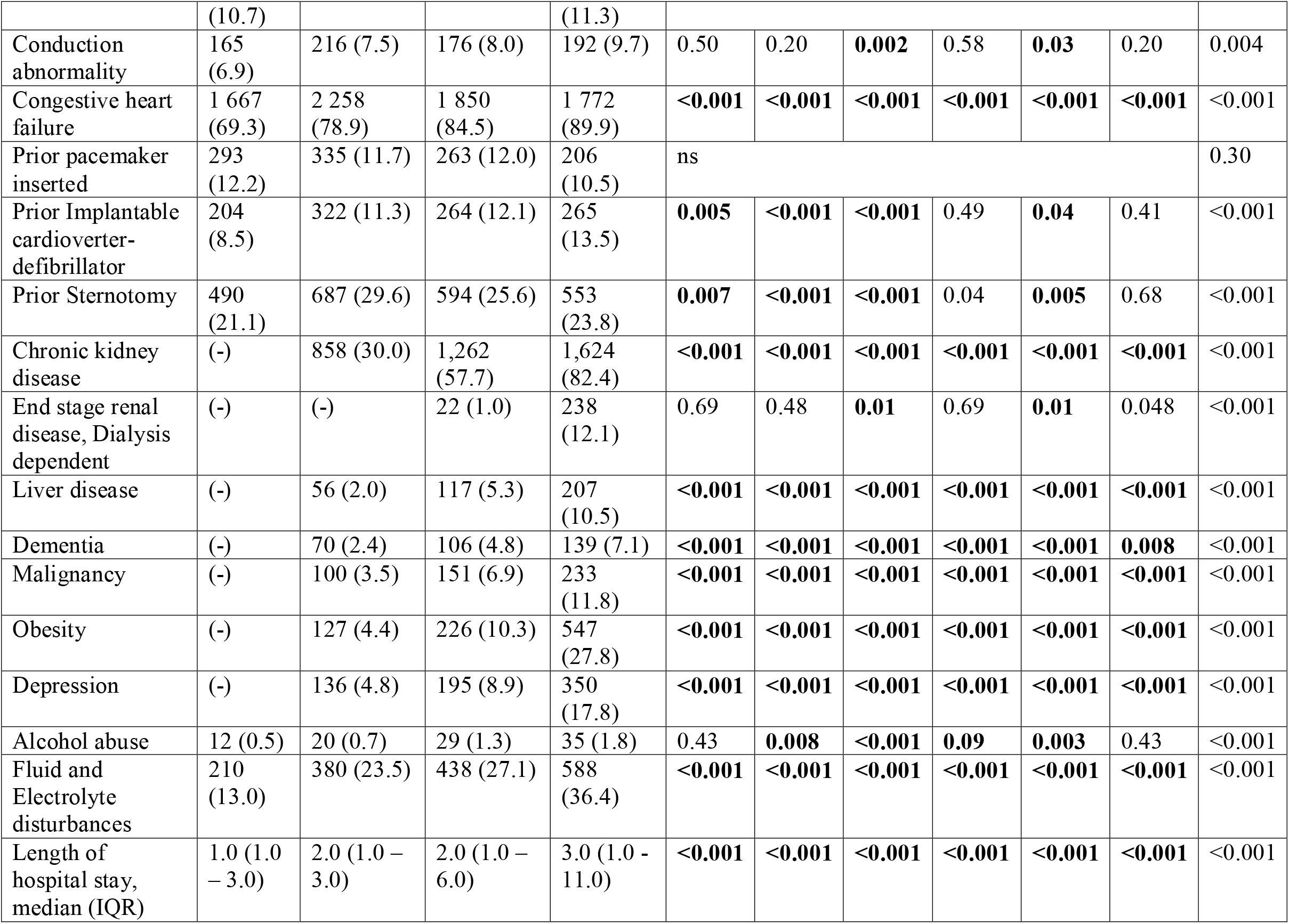
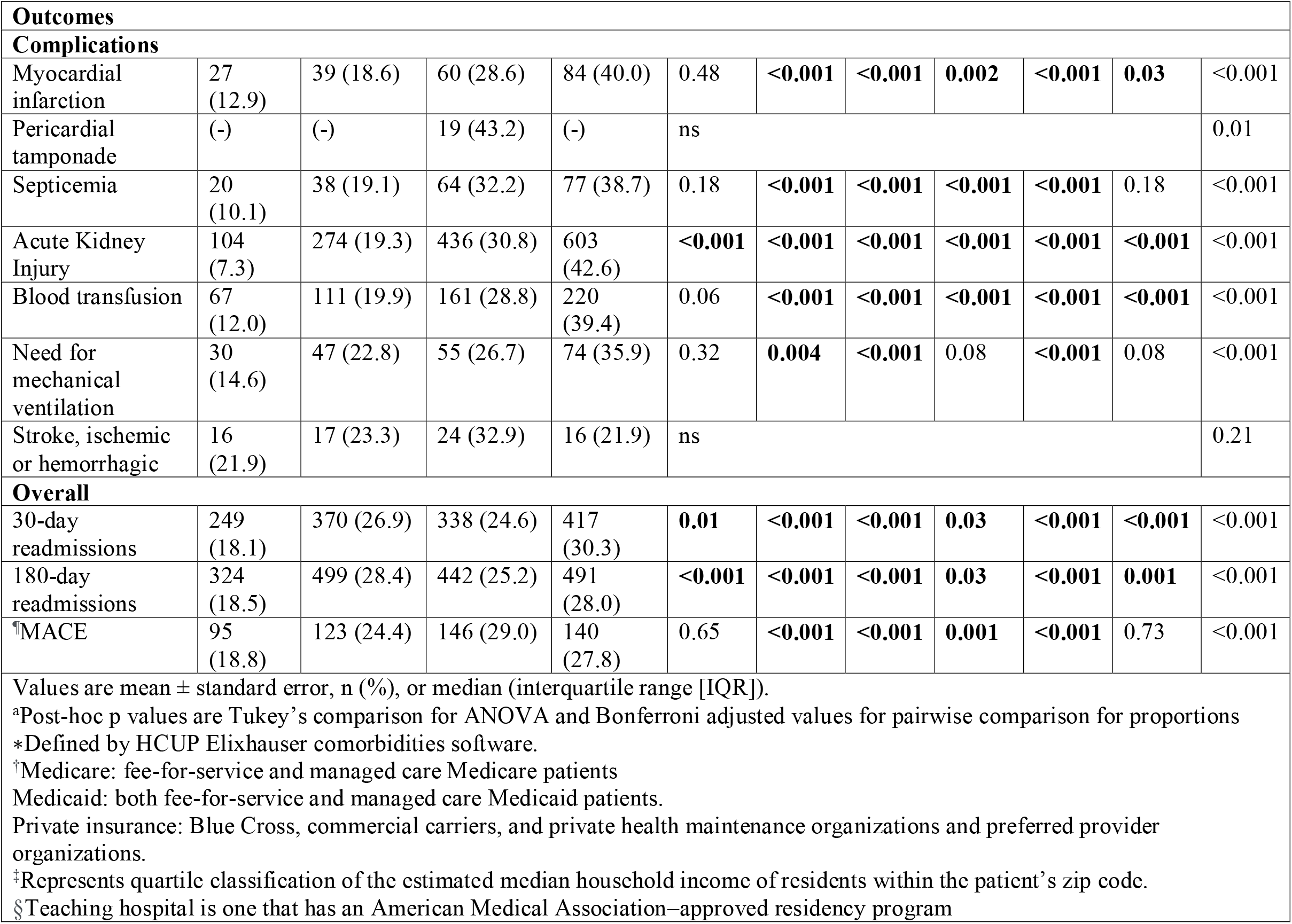
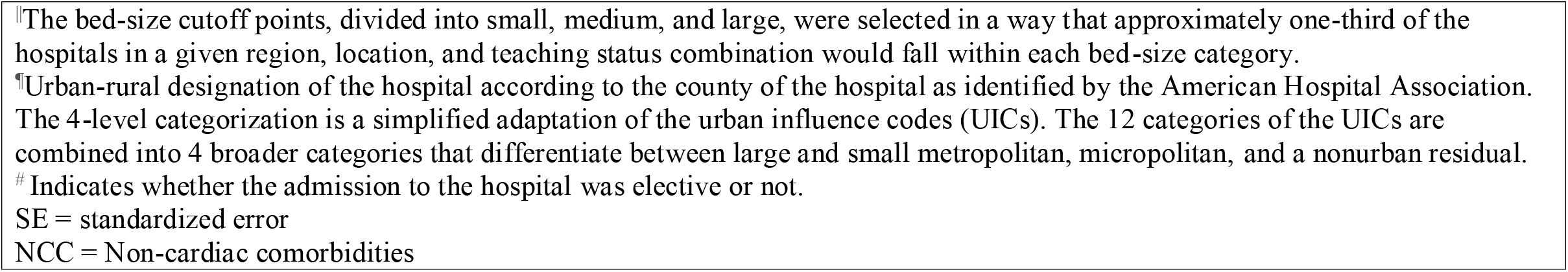
Baseline Individual- and Hospital-Level Characteristics for M-TEER admissions Stratified by Number of Non-cardiac Comorbidities

Admissions with one NCC had more proportion of patients with hypertension (*P* < 0.05), peripheral vascular disease (*P* < 0.05), coronary artery disease (*P* < 0.05), congestive heart failure (*P* < 0.05), and prior sternotomy (*P* < 0.05). Admissions with atleast three NCCs had the highest proportion of documented fluid and electrolyte disturbances, chronic kidney disease, liver disease (excluding cirrhosis), dementia, malignancy, obesity, and depression. In contrast, admissions with no NCCs had none of these comorbidities mentioned above (*P* < 0.001 for all individual pairwise Tukey). The median length of hospital stay was the highest in admissions with atleast three NCCs [3.0 days, IQR (1.0 – 11.0)] and lowest in admissions with no NCCs [1.0 days, IQR (1.0 – 3.0)]. Patients with atleast three NCCs had the highest proportion of documented myocardial infarction, acute kidney injury, and blood transfusion (all *P* < 0.05) in comparison with other categories. 30-day readmissions were found to be the highest in patients with atleast three NCCs, whereas 180-day readmissions were found to be highest in patients with one NCC. Post hoc Tukey pairwise comparison between groups was significant (all *P* < 0.05).

### PARTIAL DEPENDENCE PLOT (PDP) OF INTERACTIONS BETWEEN NCC AND TOP MACE PREDICTORS

GB classifier shows an incremental and almost linear dependence on the number of NCCs till three NCCs but then shows no/ minimal dependence with a negative slope after that (Figure 2).

**Figure 2.**
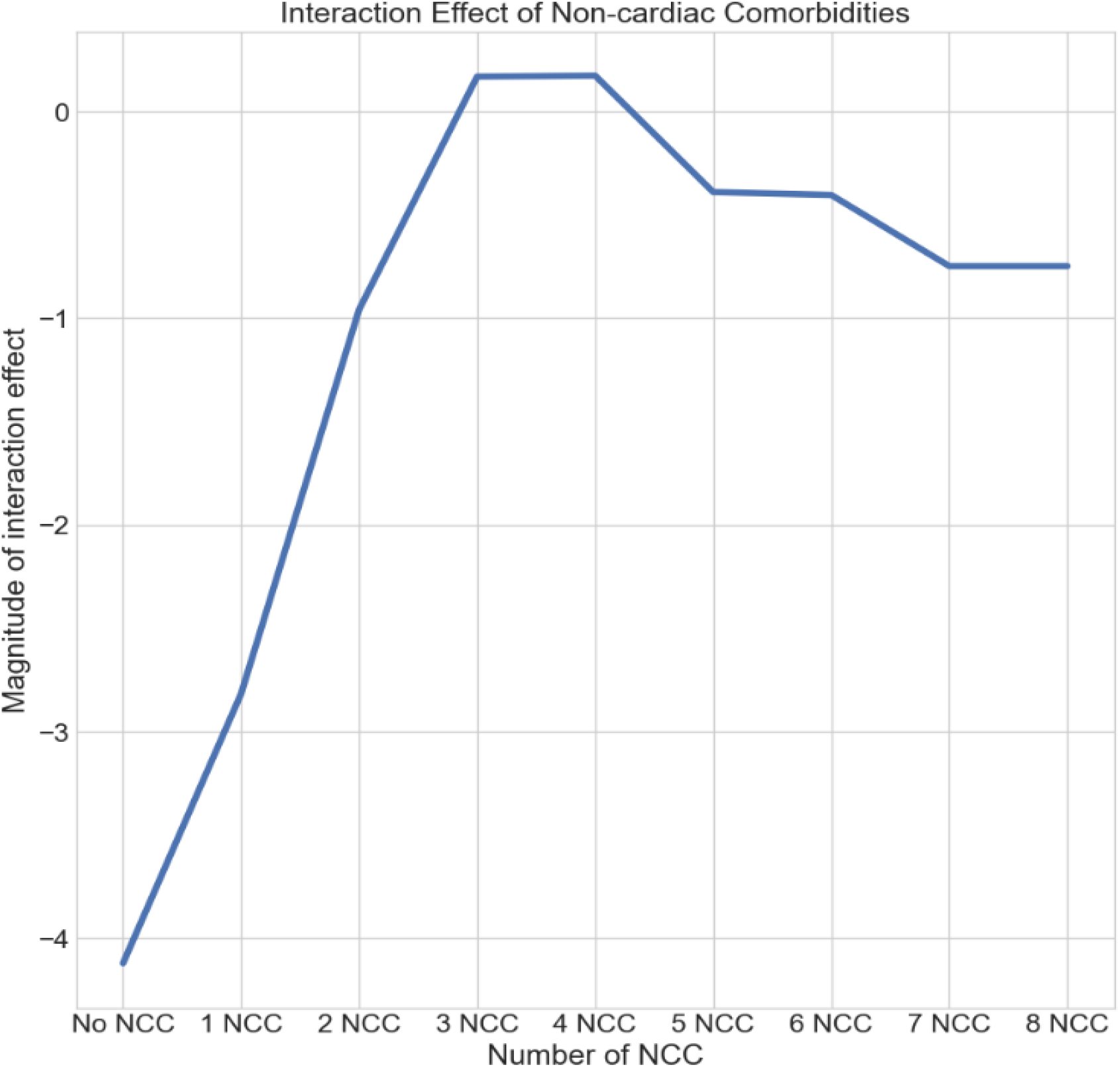
Two-Way partial dependence plot showing Interaction Effects of the Number of NCCs on MACE trained by GB Classifier. Values on the scale represent the magnitude of interaction effect. NCC=Non-cardiac comorbidities.

## DISCUSSION

The key findings in our study are 1. M-TEER procedure is safe with 504 (5.3%) in-hospital MACE out of 9 245 index admissions, 2. The median age of patients was highest in admissions with no NCC and lowest in admissions with atleast 3 NCCs, 3. Admissions with no NCCs have a higher proportion of females, 4. Most of the admissions with no or one NCC happened in high-volume centers, whereas the majority of patients with two NCCs were admitted in low-volume centers, 3. The median cost was the highest in admissions with atleast three NCCs amounting to $47 275.0 and lowest in admissions with no NCCs amounting to $39 374.3, 4. Patients from central and fringe Counties have higher in-hospital MACE events, 4. Patients with one NCC have the highest proportion of cardiac comorbidities when compared to other categories, 5. Patients with atleast three NCCs have the highest proportion of sick patients with the highest mean Elixhauser score, and comorbidities, 6. Gradient Boosting (GB) classifier has the best predictive performance amongst the trained ML classifiers with an AUROC of 0.96 (95% CI: 0.95 - 0.97) and 7. Non-elective admission is the topmost predictor for in-hospital MACE in M-TEER patients, followed by the number of NCCs and age, and 8. GB reveals non-linear relationships between the number of NCCs and other features in a better way than other trained models.

In our study, in-hospital MACE was recorded in 5.3% of M-TEER admissions. As per our results, admissions with atleast two NCCs have higher in-hospital MACE rates than the ones with less than two NCCs. Admissions with two and atleast three NCCs have comparable MACE rates. Meanwhile, the Gradient Boosting classifier showed the highest MACE rates in admissions with three or four NCCs, and afterward, the effect is the same as any admission that has two NCCs. This finding was comparable to an analysis of the German TRAMI registry that showed 2.7% of patients had in-hospital MACE. However, they failed to show a difference in in-hosptial MACCE rates between patients with multiple (at least two) and less than two NCCs. This finding might be due to the fact that there were only twenty one MACCE events in their whole cohort. They also found similar age groups between the NCC categories.

This is in contrast to our findings where we identified that patients with an increased number of noncardiovascular comorbidities were younger, reflecting our study population; older patients with multiple noncardiovascular comorbidities might have an increased competing risk of death and thereby less likely to be admitted in hospital for M-TEER. Nonagenarians are at the highest risk, followed by septuagenarians as identified by the Gradient Boosting classifier. A higher proportion of admissions were males with atleast one NCC. This finding was similar to the German TRAMI registry analysis results. Though, this feature was not included in the ML analysis by the Boruta algorithm implying its importance in governing in-hospital MACE outcome. A statistical analysis done on NRD showed no difference in in-hospital mortality and major complications among women compared with men admitted for TMVr.^24^ This was in concordance with the results of Boruta RF analysis. Though the number of NCCs is higher in admissions with atleast three NCCs, MACE outcome is comparable to admission having only two NCCs (Post-hoc chi-square *P* = 0.73). This discrepancy could be explained by the GB classifier predictions in two ways: Firstly, SHAP analysis ranked hospital volume as a top feature of importance. Admissions in low – medium volume centers mostly had two NCCs. This might buttress the phenomenon of volume-outcome relationship (VOR) for interventional procedures, including M-TEER. Chhatriwalla et al. addressed VOR by examining the relationship between institutional MitraClip case volume and in-hospital outcomes from 12 334 consecutive patients treated at two hundred and seventy five U.S. hospitals. Though in-hospital rates of death, stroke, and complications were infrequent, even after multivariate adjustment, they were not volume related.^25^ This is in strong contrast to what we have observed. In their analysis, the median number of MitraClip cases per site in the Transcatheter Valve Therapy (TVT) Registry was only 30, and 83 of 275 sites (30%) had sufficient experience to contribute to the third volume tertile. This puts the sample size of high-volume centers to a lower side, skewing the results and thereby masking VOR. Admissions in low-volume centers with three NCCs had the highest MACE rate, whereas the ones in high-volume centers with no NCC had the lowest MACE rate when the interaction effect of hospital volume on MACE is taken into account. Another reason why there could be a higher proportion of MACE encountered in admissions with NCC > two is that, a higher proportion of admissions had documented fluid and electrolyte disturbances. GB classifier ranked the same variable as one of the top predictors of MACE. Admissions with 2 NCCs have a higher number of patients with electrolyte disturbances than the ones with atleast three NCCs (Post hoc chi-square *P* < 0.001). MACE outcome had higher partial dependence on non-elective admission with four NCCs in comparison to elective admission with no NCC. A similar finding was shown in an analysis from a single-center retrospective series where it was shown that urgent/emergent TMVr in high-risk patients was associated with short-term mortality. Chronic lung disease was the most significant non-cardiac comorbidity to predict a positive MACE outcome as identified by the Gradient Boosting algorithm. Chronic obstructive pulmonary disease has been identified as a predictor of excess mortality in studies such as the American College of Cardiology/Society of Thoracic Surgeons TVT Registry as well as in the EVEREST II trial.^10, 29^ We observed significant differences in total charges for hospital stay, with admissions having atleast three NCCs having higher charges than the rest. This might be accounted for by the burden of NCC on patients, though length of stay could have a contributory role as well. A greater proportion of patients (> 71 years of age, as shown in Table 1) with higher Elixhauser score and longer mean length of stays had atleast three NCCs. This could be explained by the burden of NCCs. Higher proportion of admissions with atleast two NCCs was discharged to SNF in comparison with the other categories. This might be because a higher proportion of sick patients with a high mean elixhauser comorbidity index require post-procedural care in SNF.

This is the first study, to the best of our knowledge, to identify microclusters of admissions in patients undergoing M-TEER that have different in-hospital MACE outcomes taking NCCs into consideration. Our study represents an important step forward toward a better and more objective estimation of the effect of NCC on MACE outcomes in patients who undergo M-TEER.

The GB classifier is an explainable ML model that provides an intuitive understanding of the interaction between NCCs and cardiac comorbidities and the predictors of MACE in patients undergoing M-TEER. It has identified distinct patient subpopulations that have different non-cardiac comorbidity burden and corresponding MACE outcomes. Machine learning helped to identify such population groups by its distinct ability to uncover non-linear relationships between features.

## LIMITATIONS

Firstly, our findings do not establish causation. Results must be considered hypothesis-generating and it should be noted that residual measured or unmeasured confounding might have influenced these findings. As NRD is a database of linked inpatient discharge records, we were not able to capture mortality outside of hospitalizations. Also, the use of ICD codes for administrative data analysis is subject to potential error. It is important to take into account that classifier models are developed in populations, not individuals. Although the classifier models may inform practitioners regarding the estimated likelihood of complications in a patient, they cannot explain the individual variations in the influences not ascertained in clinical practice. Furthermore, future studies should determine the potential usefulness of incorporating NCC burden measure in ML models to more accurately predict factors that govern MACE outcomes in this specific population of patients.

## CONCLUSION

Calibrated gradient boosting classifier facilitates the understanding of predictors of in-hospital MACE in patients who undergo M-TEER. The number of non-cardiovascular comorbidities is a top predictor of in-hospital MACE and patients with three or four non-cardiovascular comorbidities are at the highest risk for in-hospital MACE.

## Data Availability

The US Healthcare Cost and Utilization Project's Nationwide Readmissions Database is a publicly available dataset that can be accessed at https://hcup-us.ahrq.gov/

## Ethical approval statement

Ethical approval was not required.

## Acknowledgement

This study was possible with a generous gift from Jennifer and Robert McNeil. The funders had no role in the design and conduct of the study, in the collection, analysis, and interpretation of the data, and in the preparation, review, or approval of the manuscript.

